# Artificial Intelligence quantified prostate specific membrane antigen imaging in metastatic castrate-resistant prostate cancer patients treated with Lutetium-177-PSMA-617

**DOI:** 10.1101/2025.08.07.25333267

**Authors:** Stephen L. Yu, Xiaofei Wang, Sinjin Wen, Seth D. Holler, Marra O. Bodkin, Joanna A. Kolodney, Sidra Najeeb, Thomas F. Hogan

## Abstract

**PURPOSE:** The VISION study^1^ found that Lutetium-177 (^177^Lu)–PSMA-617 (“Lu-177”) improved overall survival in metastatic castrate resistant prostate cancer (mCRPC). We assessed whether artificial intelligence enhanced PSMA imaging in mCRPC patients starting Lu-177 could identify those with better treatment outcomes.

**PATIENTS AND METHODS:** We conducted a single site, tertiary center, retrospective cohort study in 51 consecutive mCRPC patients treated 2022-2024 with Lu-177. These patients had received most standard treatments, with disease progression. Planned treatment was Lu-177 every 6 weeks while continuing androgen deprivation therapy. Before starting treatment, PSMA images were analyzed for SUVmax and quantified tumor volume using artificial intelligence software (aPROMISE, Exinni Inc.).

**RESULTS:** Fifty-one mCRPC patients were treated with Lu-177; 33 (65%) received 4 or more treatment cycles and these 33 had Kaplan-Meier median overall survival (OS) of 19.3 months and 23 (70%) surviving at 24 month data analysis.

At first cycle Lu-177, these 33 had significantly more favorable levels of serum albumin, alkaline phosphatase, calcium, glucose, prostate specific antigen (PSA), ECOG performance status, and F18 PSMA imaging SUV-maximum values – reflecting PSMA “target expression”.

In a “protocol-eligibility” analysis, 30 of the 51 patients (59%) were considered “protocol-eligible” and 21 (41%) “protocol-ineligible” based on initial clinical parameters, as defined in Methods. “Protocol-eligible” patients had OS of 14.6 mo and 63% survival at 24 months. AI-enhanced F18 PSMA quantified imaging found “protocol-eligible” tumor volume in mL to be only 39% of the volume in “ineligible” patients.

**CONCLUSION:** In this cohort of mCRPC patients receiving Lu-177, pre-treatment AI-assisted F18 PSMA imaging finding higher PSMA SUV / lower tumor volume associated with the patient’s ability to have four or more treatment cycles, protocol eligibility, and better overall survival.

**KEY POINTS:** *Question:* In mCRPC patients initiating Lu-177 therapy, can AI-assisted F18 PSMA imaging identify patients who have better treatment outomes?

*Findings:* 33 (65%) of a 51 consecutive patient mCRPC cohort were able to receive 4 or more cycles Lu-177. These patients had significantly more favorable serum albumin, alkaline phosphatase, calcium, glucose, PSA, performance status, and higher AI-PSMA scan SUV-maximum values, with a trend toward lower PSMA tumor volumes in mL. They had Kaplan-Meier median OS of 19.3 months and 70% survived at 24 months. AI-enhanced PSMA tumor volumes (mL) in “protocol eligible” patients were significantly lower - only 40% - than tumor volumes of “protocol ineligible” patients.

*Meaning:* In this cohort of mCRPC patients receiving Lu-177, pre-treatment AI-assisted F18 PSMA imaging finding higher PSMA SUV / lower tumor volume associated with the patient’s ability to have four or more treatment cycles, protocol eligibility, and better overall survival.

## INTRODUCTION

Prostate specific membrane antigen (PSMA) is expressed in prostate epithelium, renal proximal tubules, nervous system ganglia, benign salivary gland, and jejunum brush border. PSMA expression is higher in normal prostate than in other tissues, and 100-to-1000 fold greater in prostate cancer than in normal prostate. PSMA expression level has been linked to high tumor grade, nodal and distant metastases, shorter time to tumor progression, and disease relapse.^2,3^ The extracellular domain of PSMA can be imaged by radiolabeled small molecules that bind with high affinity, such as F18 PSMA imaging.^4^

Lu-177-PSMA radionuclide therapy has been tested against PSMA-expressing mCRPC.^5^ The randomized phase II ANZUP TheraP trial, comparing Lu-177 radionuclide versus cabazitaxel, found Lu-177 efficacious, with lower toxicity, and with a 2.1 month improvement in progression-free survival.^6^ The randomized VISION study^1^ confirmed that Lu-177 radionuclide therapy significantly improved overall mCRPC patient survival (OS) from 11.3 to 15.3 months. Soon after the VISION study ended, several participating centers conducted a follow-up “Expanded Access Program” (EAP) with Lu-177 in mCRPC^7^ confirming a similar OS of 15.1 months.

After Lu-177 was FDA approved in 2022, the West Virginia University Cancer Institute (WVUCI) implemented Lu-177 treatment in an initial cohort of patients, many referred, who received one or more Lu-177 cycles with follow-up through December 2024. These 51 patients had received most standard treatments and presented with few remaining treatment options. Many were treated on a compassionate basis and were not excluded for not meeting usual clinical trial parameters of organ function or performance status.

As our clinical experience with Lu-177 developed, we noted wide patient variation in treatment response and apparent clinical benefit. We decided to look at on-treatment F18 PSMA imaging (“Pylarify”) enhanced with artificial intelligence software, to see if this could help us determine which patients benefitted from treatment.

## METHODS

All patients had positive baseline F18 PSMA imaging (“Pylarify”). First treatment-cycle PSMA imaging images were analyzed for SUVmax and quantified tumor volume using artificial intelligence software (aPROMISE, Exinni Inc.). All initial images were quantified; all subsequent PSMA images were read at WVUCI and reported as disease objective response (OR), mixed-response (MR), or progressive disease (PD). Thirty one patients (61%) had mid-treatment (+/-cycle 3) and 22 (43%) had end-of-treatment (after cycle 6) F18 PSMA imaging.

The standard treatment plan was to continue androgen deprivation therapy (ADT) while administering Lu-177 every 6 weeks for up to 6 cycles, stopping only for disease progression, adverse events, or patient choice. Physical examination and 19 clinical and laboratory parameters were recorded at each Lu-177 treatment cycle.

Patient data and treatment results were compared 1) for those receiving only 1-3 cycles Lu-177 versus those receiving 4-6 cycles, and 2) for those patients meeting “protocol eligible” parameters versus those “not protocol eligible”.

To be considered “protocol eligible” at first cycle Lu-177, patients had to meet all of the following parameters: absolute lymphocyte count >1.0 x10^3/uL, absolute neutrophil count >1.5 x10^3/uL, albumin >3 g/dL, alkaline phosphatase (alk phos), aspartate transferase (AST), and alanine transferase (ALT) U/L all < 1.5x upper limits of normal, calcium between 7-11 g/dL, creatinine <2 g/dL, glucose between 70-250 g/dL, hemoglobin >8 g/dL, performance status (ECOG) 0-1, and platelet count >100,000 x10^3/uL.

Data collection and analysis was approved by the WVU Health Sciences Institutional Review Board. The recruitment period was between January 1, 2022 and December 31, 2024. On December 31, 2024 data – medical records, were accessed, and analysis was completed. Enrolled subjects had consent formed signed prior to treatment with Lu-177 and the authors diagnosed and treated patients during and after data collection. De-identified data was tabulated and reported using descriptive statistics. Fisher’s exact test and Wilcoxon rank test were used in the data analysis of categorical variables and continuous variables, respectively. Kaplan-Meier method and log-rank test were used in the data analysis of survival outcomes.

## RESULTS

These 51 patients all received at least one cycle of Lu-177. The majority were referred to WVUCI by outside oncologists or urologists. Patient home address zip codes at treatment onset showed that all 51 patients lived in Appalachia.

Patient age at time of original prostate cancer (PC) diagnosis was median 67 years (range 46-85) and age at first cycle of Lu-177 was median 76 years (range 49-94). Time from PC diagnosis to first cycle of Lu-177 therapy was a median of 5.6 years (range 0.9-31) – this wide range in time from diagnosis to therapy was thought to reflect the biologic variability of PC.

By retrospective review of any available old records, at original PC diagnosis median PSA was 33 ng/ml (N=41; range 0-6380); Gleason score was 8-10 in 31 (N=39; 79%); and disease was stage 4 at diagnosis in 35 (N=48; 73%). Thus, a significant portion of these patients had “high risk” disease years earlier, at time of original diagnosis.

When these 51 patients started Lu-177 therapy at WVUCI, 18 (35%) were ECOG performance status (PS) 2-4, and 20 (39%) had pain scores of 5-to10 (0-10 scale) and/or were taking opioid pain medication.

Patients had extensive prior systemic treatment (Table 1); although 45% had prostate or regional nodal radiation, only 18% had prostatectomy. Predominant disease sites at cycle 1 Lu-177 included bone (96%), prostate (82%), lymph nodes (70%), and viscera (16%).

**Table 1:** mCRPC Patient Parameters At First Cycle (C1) Lu-177.

| **Treatments Before C1 of Lu-177 (N, %)** |  |
| --- | --- |
| Abiraterone, Enzalutamide, Darolutamide | 50 (98%) |
| Androgen Deprivation Therapy (LHRH, Orchiectomy) | 50 (98%) |
| Taxane (Any) | 47 (92%) |
| Radiation Therapy (Non-Prostate/Regional Node) | 34 (67%) |
| Bone Agent (Zolendronate, Denosumab) | 25 (54%) |
| Bicalutamide, Flutamide | 27 (53%) |
| Radiation Therapy (Prostate/Regional Node) | 23 (45%) |
| Radiation Therapy with Ra-223 | 3 (6%) |
| Immunotherapy | 1 (2%) |
| Tyrosine Kinase Inhibitor (TKI) | 0 (0%) |
| **Disease Sites at C1 of Lu-177 (N, %)** |  |
| Bone | 49 (96%) |
| Prostate present | 42 (82%) |
| Nodes (Any) | 35 (70%) |
| Nodes, Extra-pelvic | 34 (65%) |
| Nodes, Pelvic | 33 (65%) |
| Total Visceral (Adrenal, Liver, Lung) | 8 (16%) |
| Liver | 6 (12%) |
| Soft Tissue (other) | 7 (14%) |
| Lung | 5 (10%) |
| **Performance Status and Pain at C1 of Lu-177 (N/Total, (%))** |  |
| ECOG 0-1 | 31/46 (67%) |
| ECOG 2-4 | 15/46 (33%) |
| Pain Scale 5/10 or More, or Narcotics Use | 18/50 (36%) |
| **Laboratory Values at C1 of Lu-177 (Median, (Range))** |  |
| Absolute Lymphocyte Count (ALC) x 10<sup>3</sup>/μL | 0.88 (0.20-3.20) |
| Absolute Neutrophil Count (ANC) x 10<sup>3</sup>/μL | 4.23 (1.8-8.96) |
| Alanine Aminotransferase (U/L) | 13 (5-107) |
| Albumin (mg/dL) | 3.4 (2.4-4.4) |
| Alkaline phosphatase (U/L) | 96.0 (3.1-1714.0) |
| Aspartate Aminotransferase (U/L) | 20 (9-72) |
| Body Mass Index (kg/m<sup>2</sup>) | 27.2 (20.5-46.2) |
| Calcium (mg/dL) | 8.85 (6.90-10.00) |
| Creatinine (mg/dL) | 0.88 (0.60-2.21) |
| Glucose (mg/dL) | 105.5 (44.0-339.0) |
| Hemoglobin (g/dL) | 10.7 (6.6-14.9) |
| Platelet Count x 10<sup>3</sup>/μL | 235 (74-591) |
| Prostate Specific Antigen (ng/mL) | 57.55 (0.90-3338.00) |
| RBC Mean Corpuscular Volume (fL) | 90.9 (80.8-103.0) |

Clinical parameters recorded at cycle 1 Lu-177 had wide ranges, and 21 (41%) would not have met usual clinical trial on-study standards (see Table 1). As just one example, cycle 1 median lymphocyte count (0.88 x 10^3^/microL) indicated that many patients were lymphopenic when starting Lu-177.

Treatment outcomes for this 51 patient cohort are shown in Table 2. This cohort had 216 cycles of Lu-177, averaging 4.2 cycles per patient. During Lu-177 therapy, 50%-PSA decline occurred in 42%; ECOG PS improved in 23%; pain scores improved to level 0-4 in 22%.

**Table 2:** mCRPC Patient Treatment Outcomes After First Cycle (C1) Lu-177.

| Clinical Outcomes |  |  |
| --- | --- | --- |
| Total Cycles | 216 |  |
| Average # Cycles/Patient | 4.2 |  |
| PSA 50% Decline during treatment (N/Total,%) | 21/50, 42% |  |
| ECOG PS Improved 1 or More (N/Total,%) | 10/44, 23% |  |
| Pain Improved to Below 5/10 (N/Total, %) | 10/45, 22% |  |
| Post-Lu-177 PSMA-PET Imaging Results | Cycle 3 | Cycle 6 or End of Treatment |
| Patients Imaged (N) | 31 | 22 |
| OR | 13 (42%) | 8 (36%) |
| MR | 8 (26%) | 5 (23%) |
| PD | 10 (32%) | 9 (41%) |
| Survival Post Cycle C1 of Lu-177 |  |  |
| Median Survival and Range (in months) | 14 (9.4-25+) |  |
| Patients Alive as of December 2024 (N, %) | 25 (49%) |  |

In the subset of 31 patients with “mid-treatment” PSMA imaging, usually just after cycle 3, OR was reported in 42%, MR in 26%, and PD in 32%; in the subset of 22 patients with “end-of-treatment” PSMA imaging, responses were similar, with OR 36%, MR in 23%, and PD in 41%.

The Kaplan-Meier median overall survival for the entire cohort was 14 months (9.4-25+), and 25 of the 51 patients (49%) remained alive at the 24 month data analysis December 2024 (Figure 1a; Table 2).

**Figure 1:**
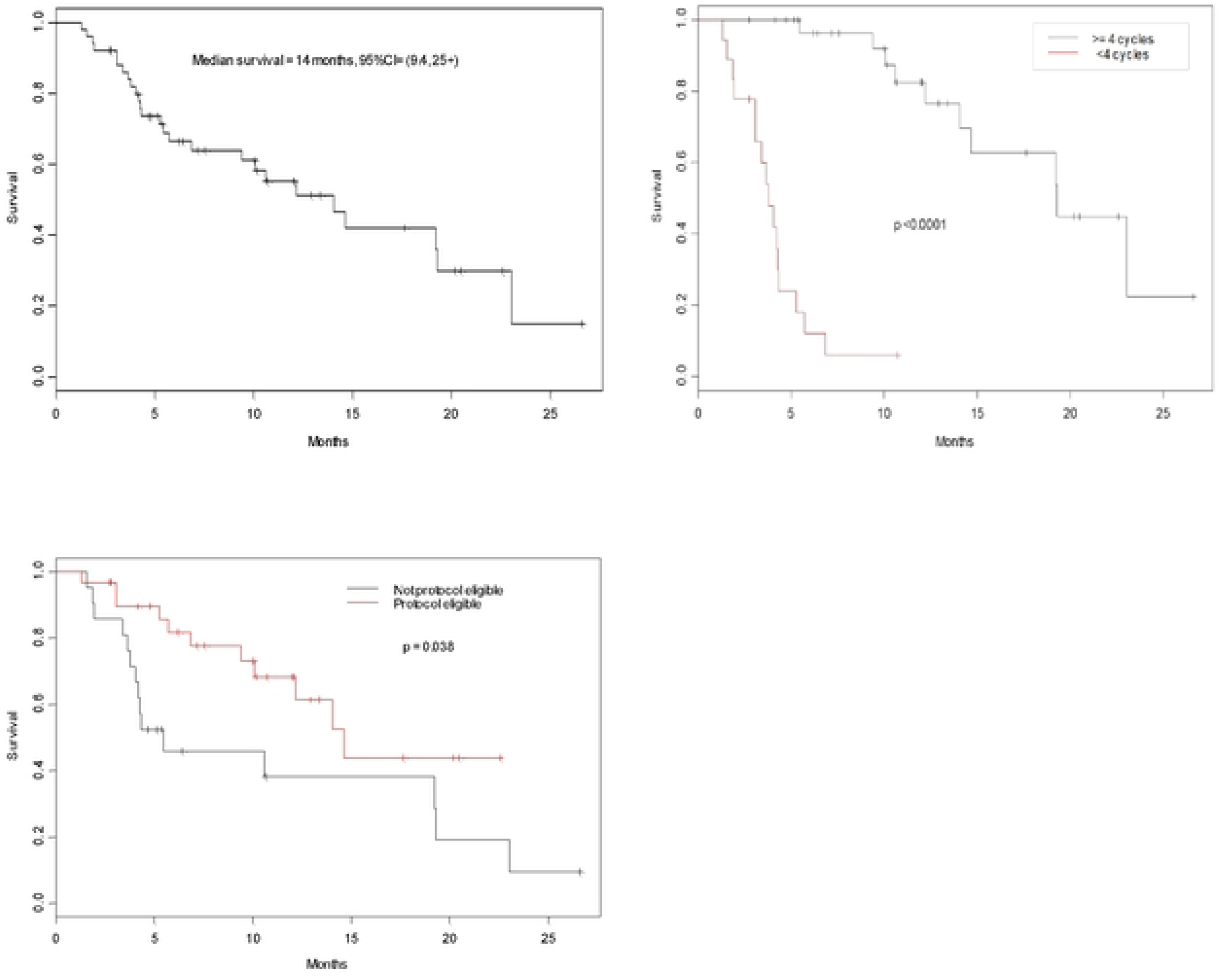
Median months’ survival after starting Lu-177 (177-Lu PSMA-617) therapy in 51 mCRPC patients. A) entire group (14 mo) B) subgroups 1-3 cycles (3.8 mo) versus 4 or more cycles (19.3 mo) C) subgroups “protocol not-eligible” (5.5 mo) versus “protocol eligible” (14.6 mo)

To further study the utility of Lu-177 in this 51 patient mCRPC cohort, we subdivided the patients two different ways, comparing outcomes for each subset.

First, we looked at the 33 (65%) patients receiving 4 or more cycles Lu-177 versus the 18 (35%) patients receiving 3 or fewer cycles. Comparing these two groups, we found no significant differences at start of Lu-177 therapy in patient age, ALT, AST, BMI, cholesterol, creatinine, number of disease sites (bone, liver, lung, lymph nodes), platelet count, time from 1^st^ diagnosis to treatment, or white blood count.

However, the groups appeared significantly different at treatment onset in levels of serum albumin, alkaline phosphatase, calcium, glucose, PSA, ECOG PS, AI-PSMA imaging SUV-maximum values, and in risk of deaths (Table 3). Also, there was a non-significant trend in those receiving 4 or more cycles having lower AI-PSMA imaging quantified tumor volumes in mL. Patients receiving 4 or more Lu-177 cycles had Kaplan-Meier median OS of 19.3 months with 23 of 33 (70%) surviving at 24 month data analysis. In contrast, patients receiving only 1-3 Lu-177 cycles had Kaplan-Meier median OS of 3.8 months with only 2 of 18 (11%) surviving at 24 month data analysis (Figure 1b; Table 3)

**Table 3:** mCRPC Patient Comparison: 1-3 Versus 4 Or More Cycles Lu-177.

|  | **Cycle 1-3** | **Cycle 4 or More** | **P-value** |
| --- | --- | --- | --- |
| **Number of patients (%)** | 18 (35%) | 33 (65%) |  |
| **Patient Demographics** |  |  |  |
| Median Age at C1 of Lu-177 (yrs) | 72.9 | 77.6 | NS |
| Median Overall Survival, months (range) | 3.8 (1.5-17.7) | 19.3 (1.3-26.6) | 0.001 |
| Patients Alive as of December 2024 (N, %) | 2 (11%) | 23 (70%) | 0.001 |
| **Treatment Before Cycle 1 Lu-177 (N, %)** | **Cycle 1-3** | **Cycle 4 or More** | **P-value** |
| Bone Agent – Zoledronate, Denosumab | 6 (33%) | 19 (58%) | NS |
| Radiation (prostate/regional nodes) | 13 (72%) | 21 (64%) | NS |
| Radiation (not to prostate/regional nodes) | 7 (39%) | 16 (48%) | NS |
| Prostatectomy | 5 (28%) | 4 (12%) | NS |
| **Clinical Data at Cycle 1 of Lu-177** | **Cycle 1-3** | **Cycle 4 or More** | **P-value** |
| Median Body Mass Index (BMI) | 28.0 | 26.5 | NS |
| Performance Status 0-1 | 8 (17%) | 25 (54%) | 0.041 |
| Pain Score 5 or more | 9 (18%) | 10 (20%) | NS |
| **Median Lab Values at Cycle 1 of Lu-177** | **Cycle 1-3** | **Cycle 4 or More** | **P-value** |
| Absolute Lymphocyte Count (ALC) x 10<sup>3</sup>/μL | 0.7 | 0.9 | NS |
| Albumin (mg/dL) | 3.3 | 3.5 | 0.038 |
| Alkaline phosphatase (U/L) | 120.5 | 86.0 | 0.065 |
| Calcium (mg/dL) | 8.6 | 9.1 | 0.003 |
| Creatinine (mg/dL) | 1.0 | 0.8 | 0.068 |
| Glucose (mg/dL) | 94.5 | 115 | 0.044 |
| Prostate Specific Antigen (ng/mL) | 101.5 | 28.7 | 0.006 |
| AI-PSMA Imaging SUVmax, mean (SD) | 52 (43.1) | 92.9 (64.77 ) | 0.017 |
| AI-PSMA Imaging Tumor Vol, mean (SD) | 957 (1367) | 598 (709) | NS trend |

Second, we subdivided the 51 patient cohort into 30 patients (59%) meeting “protocol eligibility parameters” -- as defined in the methods section -- versus 21 patients (41%) not meeting these “protocol eligibility”. Comparing these two groups, we found no significant differences when starting Lu-177 therapy in patient age, levels of ALT, AST, BMI, cholesterol, creatinine, glucose, WBC, platelets, or number of disease sites (bone, liver, lung, lymph nodes).

However, the “protocol eligible” patients had significantly better levels of albumin, alkaline phosphatase, calcium, ECOG PS, hemoglobin, PSA, risk of death, and AI-PSMA imaging quantified tumor volumes in mL. Using artificial intelligence software (aPROMISE, Exinni Inc.) found the tumor volume in the “protocol eligible” group to be 39% (p 0.003) of the “protocol ineligible” group (Table 4). The 30 “protocol eligible” patients had Kaplan-Meier median OS of 14.6 months with 19 (63%) surviving at data analysis. (Figure 1c; Table 4).

**Table 4:** mCRPC Patient outcomes by “protocol eligibility” as defined in methods.

|  | **Not Eligible** | **Eligible** | **P-value** |
| --- | --- | --- | --- |
| Patients (N/51 total, %) | 21 (41%) | 30 (59%) |  |
| Patients Getting Only 1-3 Cycles of Lu-177 | 10/21 (48%) | 8/30 (27%) | NS trend |
| Median Age at C1 of Lu-177 (yrs) | 73.9 | 77.0 | NS |
| Median Time, Diagnosis to Lu-177 (yrs) | 4.7 | 8.2 | NS |
| Median Overall Survival (mos (range)) | 5.5 (1.5-26.6) | 14.6 (1.3-20.6) | 0.038 |
| Patients Alive as of Dec 2024 (N, %) | 6/21 (29%) | 19/30 (63%) | 0.023 |
| **Selected Treatments pre-Cycle 1 Lu-177 (# Pts, %)** | **Not Eligible** | **Eligible** | **P-value** |
| Radiation (prostate/regional nodes) | 10 (48%) | 13 (43%) | NS |
| Radiation (other) | 16 (76%) | 18 (60%) | NS |
| Prostatectomy | 2 (10%) | 7 (23%) | NS |
| **Clinical Data at Cycle 1 of Lu-177** | **Not Eligible** | **Eligible** | **P-value** |
| Median BMI (kg/m<sup>2</sup>) | 27.4 | 27.2 | NS |
| Performance Status 0-1 (N) | 9 | 24 | 0.034 |
| **Median Lab Values at Cycle 1 Lu-177** | **Not Eligible** | **Eligible** | **P-value** |
| Absolute Lymphocyte Count (ALC) x 10<sup>3</sup>/μL | 0.7 | 1.1 | 0.01 |
| Albumin (mg/dL) | 3.3 | 3.5 | 0.039 |
| Alkaline phosphatase (U/L) | 198.0 | 83.5 | 0.004 |
| Calcium (mg/dL) | 8.8 | 9.1 | 0.059 |
| Hemoglobin (g/dL) | 10.0 | 11.7 | 0.007 |
| Prostate Specific Antigen (ng/mL) | 90.5 | 33.8 | 0.043 |
| AI-PSMA Imaging SUVmax, mean (SD) | 68.4 (50.9) | 86.7 (66.8) | NS |
| AI-PSMA Imaging Tumor mL, mean (SD) | 1131 (1180) | 448 (714) | 0.003 |

Of interest, in both of the above analyses, patients with higher SUVmax values - reflecting PSMA target expression - and with lower AI-PSMA imaging quantified tumor volumes in mL, were those able to have four or more treatment cycles, were more protocol eligibile, and had better overall survival. (Tables 3,4)

## DISCUSSION

Metastatic castration-resistant prostate cancer (mCRPC) is lethal, globally resulting in over 375,000 deaths annually.^8^ The Lancet Commission on prostate cancer expects an impending worldwide surge in the number of cases,^9^ and improving the benefit of current PC treatments will be necessary.

Our initial experience at WVUCI found that our mCRPC patients selected for Lutetium-177-PSMA-617 therapy -- many referred -- presented with advanced disease, prior extensive treatment, and few remaining treatment options. Many of these Appalachian patients had inferior clinical and laboratory parameters at treatment onset, versus the landmark VISION clinical trial patients.^1^ As just one example, many were lymphopenic at treatment onset -- a known risk factor associated with poor treatment outcomes in other solid tumors.^10,11^ For the 51 patients reported here, Lu-177 was expensive, with estimated isotope-only cost of $231,000 per patient, a portion of which was affordable only with generous manufacturer subsidies.

From our initial patient experience, we see that we need to improve patient selection for Lu-177 therapy, and to apply this treatment in a different context. Currently, ideas to improve mCRPC patient outcomes include using Lu-177-PSMA-617 earlier in the prostate cancer treatment space, prior to systemic chemotherapy or palliative radiation.^12^

Although not part of our original hypothesis, we found that getting F18 PSMA re-imaging (“Pylarify”) after only three treatment cycles seemed to correspond to later results, identifying early treatment success or failure (Table 2). This impression can be further validated as we accrue a larger number of patients.

The aPROMISE software used for automated image analysis of PSMA PET/CT was FDA-cleared in 2021. ^13^ The software offers quantitative analysis of hotspots and standardized reporting of PSMA PET/CT scans. Sensitivity for detecting suspicious metastases was 87% (bone) and 92% (regional lymph nodes), with a number of false positives, but high inter-reader agreement. ^14,15^

Potential issues with the current study include the fact that it is retrospective, from a single institution, with a small initial cohort, for a geographically restricted Appalachian region, retrospective, not excluded for treatment by protocol eligibility.

Going forward, we think that quantifying prostate cancer tumor burden via AI-enhanced PSMA imaging is very promising and deserves further study for its potential contribution to patient treatment selection. In the mCRPC patients reported here, analysis of on-treatment AI-PSMA imaging with AI-enhancement software found that patients with higher SUVmax values (reflecting PSMA expression) and with lower tumor volumes were the ones receiving four or more treatment cycles of Lu-177, were more likely to be “protocol eligible”, and had longer survival (Tables 3,4)

We encourage regional clinicians caring for mCRPC patients to consider using Lutetium-177-PSMA-617 earlier in the prostate cancer spectrum. We now intend to re-purpose Lutetium-177-PSMA-617 for our mCRPC patients prior to chemotherapy and optimize patient selection with AI-quantified PSMA imaging prior to treatment.

## Data Availability

The core data is HIPPA protected and cannot be shared publicly. De-identified data is held privately in the Dept of Oncology, West Virginia University, Morgantown, WV. The authors will honor reasonable requests to share de-identified data on a case-by-case basis.

## ACKNOWLEDGEMENTS

The authors thank Laurel Lyckholm MD, Michael Kolodney MD, PhD, and Samuel Merrill MD, PhD for helpful insights and manuscript review.

## AUTHOR CONTRIBUTIONS

Conception: SY, SH, TH

Patient Management: XW, SH, MB, JK, SN, TH Data Collection: SY, XW, SH, TH

Data Analysis: XW, SW, SH, TH

Figures-Graphics: SH, SW

Manuscript Writing: SY, SH, TH

Manuscript Approval: all authors

## ABBREVIATIONS USED

ADT: androgen deprivation therapy
AI: artificial intelligence
ALC: absolute lymphocyte count
ALT: alanine aminotransferase
ANC: absolute neutrophil count
ARPI: androgen receptor pathway inhibitors
AST: aspartate aminotransferase
BMI: body mass index (kg/m2)
CBC: complete blood counts
cfDNA: cell-free DNA
CMP: comprehensive metabolic panel
CRPC: castration resistant prostate cancer
ctDNA: circulating tumor DNA
EAP: Expanded Access Program;
ECOG: Eastern Cooperative Oncology Group
FDA: United States Food and Drug Administration
Lu-177 therapy: Lu-177-PSMA-617 (“Lu-177”) therapy for PC
MR: mixed-response
OR: objective response
OS: overall survival
PC: prostate cancer
PD: progressive disease
PS: clinical performance status (ECOG)
PSA: prostate specific antigen
PSMA: prostate specific membrane antigen PSMA
Imaging: F18 piflufolastat PET imaging of PC (“PSMA imaging”)
Pyl.BL.T.volume: Total tumor volume in mL identified by AI enhanced PSMA scans
SUVmax: maximum standardized uptake value of PSMA imagine.
WVUCI: West Virginia University Cancer Institute.

